# Diagnostic accuracy and clinician-rated reasoning quality dissociate in a blinded evaluation of large language models in psychiatry

**DOI:** 10.64898/2026.02.03.26345402

**Authors:** Kevin W. Jin, Yasna Rostam-Abadi, Pooja Chaudhary, Margaret A. Garrett, Ashley S. Huang, Mario Montelongo, Caesa Nagpal, Jasperina Shei, Judah Weathers, Juliana S. Zhang, Manu S. Sharma, Qingyu Chen, Jiyeong Kim, Matteo Malgaroli, Walter S. Mathis, Carolyn I. Rodriguez, Salih Selek, Christopher Pittenger, Sarah W. Yip, Brian A. Zaboski, Hua Xu

## Abstract

**Background:** Evaluations of large language models (LLMs) in medicine rest largely on diagnostic accuracy. We report what we learned evaluating four frontier LLMs on psychiatric diagnosis, where we asked whether clinician assessment of model reasoning adds information that accuracy does not.

**Methods:** Four LLMs produced ranked five-diagnosis differentials for 196 psychiatric vignettes (135 screened published case reports; 61 unpublished, clinician-authored). An automated adjudicator scored accuracy. On a random 30-vignette subset, five psychiatrists blinded to model identity judged correctness and rated reasoning on a 0-4 rubric adapted from the ACGME Psychiatry Milestones. We validated the adjudicator against the clinicians, measured the reproducibility of accuracy across generation runs and repeated adjudication, and tested whether computable reasoning trace features could substitute for clinician ratings.

**Results:** On all 196 vignettes, automated top-5 accuracy ranged from 0.759 to 0.837, and the majority verdict of three adjudication passes separated one of six model pairs. On the 30 rated vignettes, clinician-determined accuracy separated no pair, while reasoning separated five. Two systems were equally accurate (0.827 each; difference bounded within ±0.059, or ±0.10 after Holm adjustment) yet differed by 0.70 reasoning points (95% CI, 0.50 to 0.90). The adjudicator agreed with held-out clinician majorities less closely than individual clinicians did (0.83 to 0.85 vs 0.89 to 0.96), regeneration changed top-1 correctness for 18 of 119 pairs, and reference strings that combined several diagnoses were scored as misses until split. Trace features that tracked ratings across model averages did not track them across individual traces.

**Conclusions:** Accuracy did not distinguish models that clinicians rated differently, and automated accuracy depended on how it was scored. Where LLMs are meant to support clinical reasoning, structured clinician assessment of reasoning belongs in pre-deployment evaluation.

## Introduction

Evaluations of large language models (LLMs) in medicine rest almost entirely on diagnostic accuracy,^1–4^ and as frontier systems converge on benchmarks, accuracy discriminates less between them.^5–10^ For a model meant to support clinical reasoning, where a clinician must decide whether to accept, question, or act on its output, two equally accurate systems may not be equivalent.^11^ Contemporary models emit clinician-visible reasoning traces, which makes that reasoning available for structured assessment.^12–19^

Psychiatry, where fluent output is easily mistaken for valid clinical reasoning,^20^ represents a demanding test. Diagnosis depends on narrative interpretation rather than biomarkers, comorbidity is common, and clinicians often disagree, so the reference standard is itself less stable than in fields with definitive tests.^21–23^ In non-psychiatric domains, LLMs have been shown to perform comparably to clinicians in controlled diagnostic evaluations;^24^ however, in psychiatry, evaluations have relied on small vignette sets, narrow diagnostic scope, older models, or accuracy alone.^25–31^

We evaluated four frontier LLMs on psychiatric vignettes and investigated whether structured clinician assessment of reasoning yields information that accuracy does not, and whether computable features of the reasoning trace could substitute for it. Building the evaluation also revealed how fragile automated accuracy can be, and we report those lessons alongside the main result.

## Methods

### Dataset

We assembled 196 vignettes (**Figure 1**). Of 183 published diagnostic case reports from *Case Reports in Psychiatry, NEJM, NEJM Evidence, JAMA, JAMA Psychiatry, The Lancet*, and *DSM-5-TR Clinical Cases*, each was given a single score by one of seven psychiatrists or residents, from 1 (worst) to 5 (best), for diagnostic clarity, narrative coherence, and completeness; the 48 scoring below 3 were excluded, leaving 135, and explicit diagnostic labels were removed. Six clinicians wrote 61 further vignettes from clinical experience; these are absent from training corpora, and free of publication selection. Reference diagnoses spanned 19 DSM-5-TR classes. Vignettes were presented as written, so diagnoses with duration or course criteria depended on the history each vignette gave. Case mix, length, reference formats, and screening scores are in **Section S1**. The study used no patient data and recruited no participants. Models

**Figure 1.**
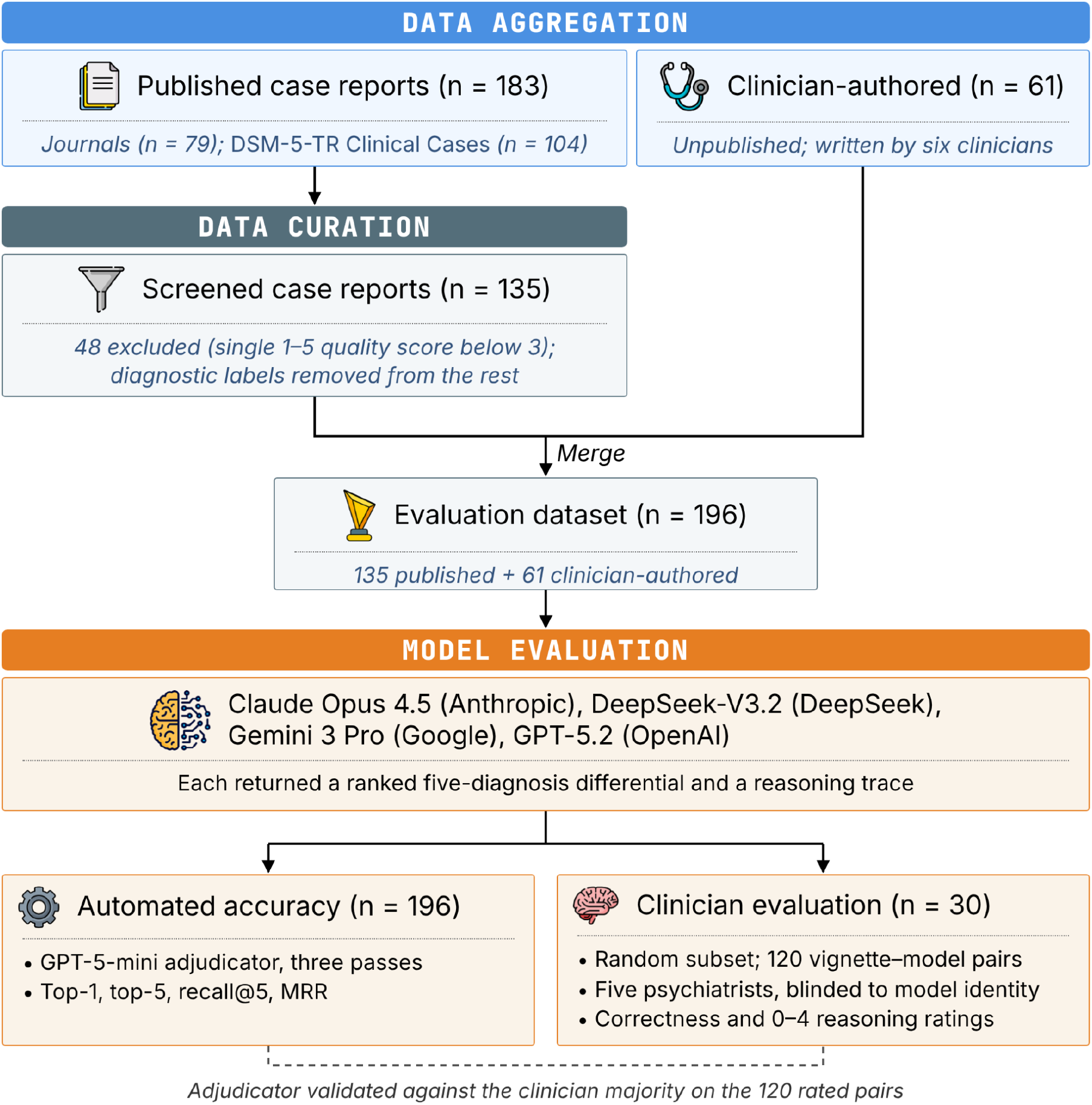
Study workflow. Each of 183 published case reports was given a single score from 1 (worst) to 5 (best) by one of seven psychiatrists or psychiatry residents for diagnostic clarity, narrative coherence, and completeness; the 48 scoring below 3 were excluded and diagnostic labels were removed from the remaining 135. With 61 clinician-authored vignettes, the final dataset comprised 196. Four models each produced a ranked five-diagnosis differential with a reasoning trace for every vignette. Accuracy was scored by an automated adjudicator on all 196; five psychiatrists, blinded to model identity, judged correctness and rated reasoning on a random 30-vignette subset, and the adjudicator was validated against their majority verdict.

Three proprietary models, Claude Opus 4.5 (claude-opus-4-5-20251101), Gemini 3 Pro (gemini-3-pro-preview), and GPT-5.2, and one open-weight model, DeepSeek-V3.2, were accessed through standard provider APIs between December 2025 and January 2026 with provider-recommended reasoning settings.^12–16,32–34^ A zero-shot prompt asked for the five most probable *DSM-5-TR* diagnoses in rank order with ICD-10 codes and supplied the same reasoning guidance, adapted from the ACGME Psychiatry Milestones,^35,36^ to every model; the reasoning evaluated is the trace each provider returned. DeepSeek-V3.2 was set to temperature 0, which does not ensure determinism.^37,38^ Prompts, settings, and prompt provenance are in **Section S2**.

### Accuracy and adjudication

A prediction was correct if it matched any reference diagnosis; we report top-1 accuracy, top-5 accuracy, recall@5, and mean reciprocal rank. Each prediction was compared with each reference by a string-similarity tier, or failing that, by GPT-5-mini (gpt-5-mini-2025-08-07) prompted as a strict psychiatric adjudicator. Because the adjudicator is not deterministic, every output was adjudicated three times; accuracy is the mean of the three passes, and models were compared with exact McNemar tests on the majority verdict, a test specified after the passes were run. Nine reference strings that combined several diagnoses were split into their components before scoring (**Section S4**).

### Clinician evaluation

Five psychiatrists, blinded to model identity, rated a random subset of 30 vignettes (120 vignette-model pairs). One vignette (1056) was written by a rater. For each pair, raters judged whether the model had “captured at least the primary diagnosis” and scored data extraction and diagnostic reasoning on a 0-4 rubric (**Section S3**). Correctness and reasoning were rated in the same session. A psychiatrist who did not rate reasoning scored each of the 30 vignettes for diagnostic ambiguity. Clinician determinations are the reference standard for the 30-vignette analyses, and we validated the adjudicator against the clinician majority (at least 3 of 5). Accuracy of two psychiatry residents on the same vignettes is reported for context in **Section S7**.

### Analysis

The unit of analysis was the vignette, which yields one rater-averaged score per model; models were compared pairwise within vignette with Holm-Bonferroni correction across the six comparisons. Because failing to reject a difference is not evidence of equivalence, accuracy equivalence was tested by two one-sided tests, reporting the smallest margin at which each pair is equivalent; the pair carrying the primary claim was chosen post hoc because its accuracies matched. Computable trace features (word count, distinct ICD-10 codes named, and self-correction phrases) were related to reasoning ratings across the four model means and across the 120 traces, with a random intercept for vignette. Reproducibility analyses were unplanned. Statistical details are in **Section S5**.

## Results

### Diagnostic accuracy across 196 vignettes

Top-5 accuracy ranged from 0.759 to 0.837 and top-1 accuracy from 0.573 to 0.668 (**Table 1**). Claude Opus 4.5 ranked first on every metric in every pass and had higher top-5 accuracy than Gemini 3 Pro (0.837 vs 0.759; Holm-adjusted *p* = 0.016). Its top-1 advantage over DeepSeek-V3.2 (0.668 vs 0.573) was significant in one of three passes but not in their majority verdict (*p* = 0.076), and in the majority verdict no other pair differed. Accuracy did not differ between published and clinician-authored vignettes (top-1 odds ratio, 1.18; 95% CI, 0.49 to 2.80; **Section S1**).

**Table 1.** Psychiatric diagnostic accuracy of large language models (n = 196; mean of three adjudication passes, bootstrap 95% CI)

|  | **Top-1 Accuracy** | **Top-5 Accuracy** | **Recall@5** | **Mean Reciprocal Rank** |
| --- | --- | --- | --- | --- |
| Claude Opus 4.5 | 0.668<br>(0.604-0.730) | 0.837<br>(0.786-0.884) | 0.754<br>(0.702-0.803) | 0.742<br>(0.688-0.791) |
| DeepSeek-V3.2 | 0.573<br>(0.507-0.643) | 0.784<br>(0.730-0.835) | 0.688<br>(0.631-0.742) | 0.667<br>(0.612-0.723) |
| GPT-5.2 | 0.628<br>(0.563-0.690) | 0.767<br>(0.711-0.821) | 0.701<br>(0.643-0.755) | 0.691<br>(0.635-0.746) |
| Gemini 3 Pro | 0.599<br>(0.532-0.663) | 0.759<br>(0.702-0.813) | 0.685<br>(0.629-0.739) | 0.667<br>(0.610-0.723) |

### Equally accurate models differed in rated reasoning

On the 30-vignette subset, clinician-determined accuracy ranged from 0.767 to 0.827 and mean reasoning scores from 2.37 to 3.60 (**Table 2**). Accuracy separated none of the six pairs, while reasoning separated five, by 0.48 to 1.23 points on the 0-4 scale (**Figure 2**). Claude Opus 4.5 and DeepSeek-V3.2 were correct equally often (0.827 each; accuracy difference bounded within ±0.059, or ±0.10 after Holm adjustment), yet differed by 0.70 reasoning points (95% CI, 0.50 to 0.90; Holm-adjusted *p* < 0.001). All five raters, working independently, ranked the four systems in the same order. Equivalence held for this pair only: comparisons involving GPT-5.2 are consistent with true accuracy differences of 15 to 20 points and are underpowered rather than equivalent (**Table S5A**).

**Table 2.** Clinician-determined diagnostic accuracy and clinician-rated reasoning quality (*n* = 30)

|  |  | Clinician-rated reasoning quality (0-4) |  |
| --- | --- | --- | --- |
| Model name | Clinician-determined accuracy | Data extraction and organization (mean ± SD) | Diagnostic reasoning and differential diagnosis (mean ± SD) |
| Claude Opus 4.5 | 0.827 | $3.67 \pm 0.38$ | $3.60 \pm 0.45$ |
| Gemini 3 Pro | 0.820 | $3.15 \pm 0.69$ | $3.12 \pm 0.60$ |
| DeepSeek-V3.2 | 0.827 | $2.97 \pm 0.61$ | $2.90 \pm 0.63$ |
| GPT-5.2 | 0.767 | $2.21 \pm 0.50$ | $2.37 \pm 0.52$ |

**Figure 2.**
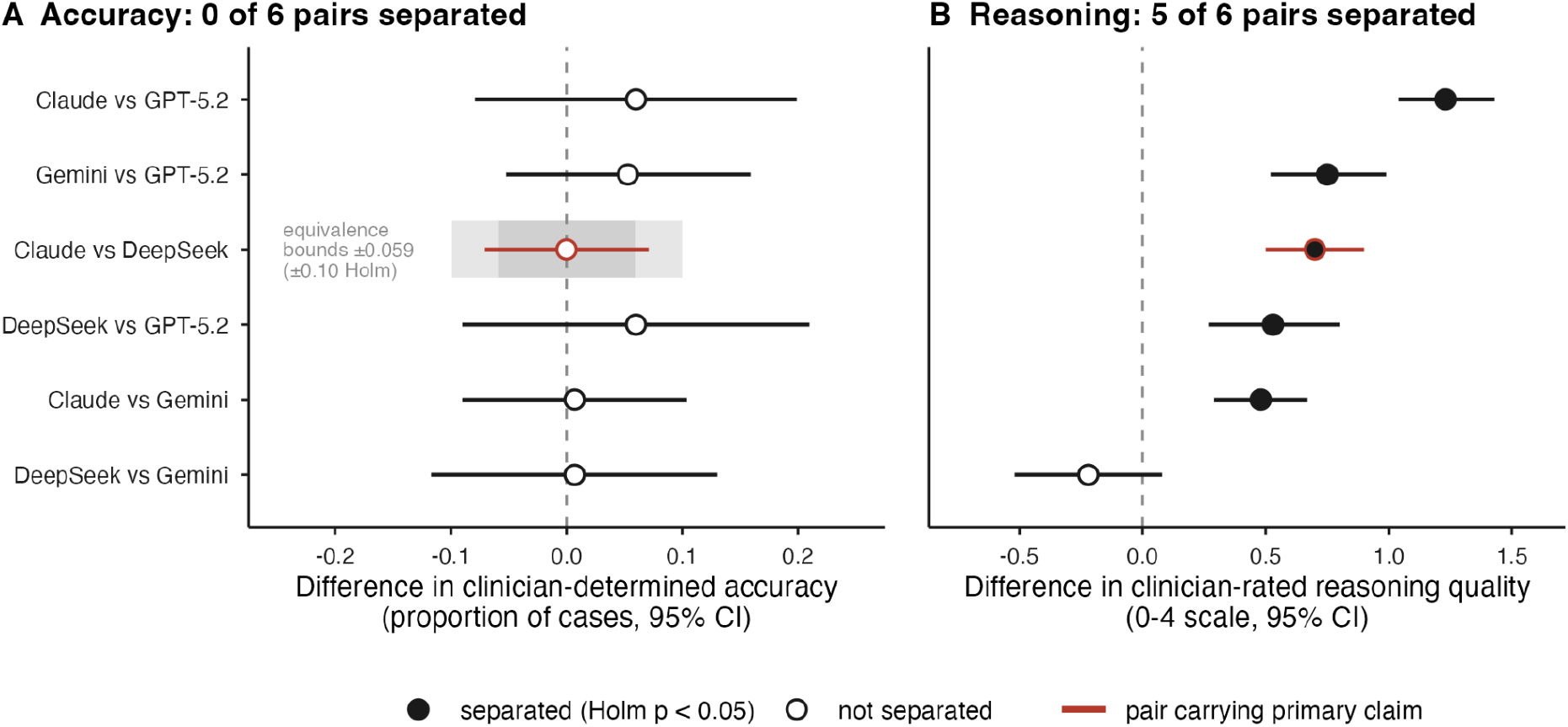
Clinician-determined diagnostic accuracy and reasoning quality dissociate (*n* = 30) Each row is one model pair; points are paired within-vignette differences (first model minus second) with 95% paired t-intervals, and filled points mark differences that remain significant after Holm adjustment across the six comparisons. **(A)** Difference in clinician-determined accuracy: for each vignette, the proportion of five psychiatrists, judging independently, who found that the model had captured at least the primary reference diagnosis in its five-item differential, averaged over vignettes. Shading on the Claude Opus 4.5 vs DeepSeek-V3.2 row marks the smallest margin at which two one-sided tests establish equivalence: ±0.059 unadjusted (dark) and ±0.10 after Holm adjustment (light). **(B)** Difference in mean diagnostic reasoning score on the 0–4 rubric, averaged over the five raters. Red marks the pair carrying the primary claim. Claude, Claude Opus 4.5; DeepSeek, DeepSeek-V3.2; Gemini, Gemini 3 Pro.

The result did not depend on ambiguous cases. The independent psychiatrist judged 23 vignettes to have a single clear diagnosis; on these, accuracy again separated no pair and reasoning the same five (**Table S5B**). Excluding the one vignette whose reference diagnosis that psychiatrist judged incorrect left every result unchanged.

Clinicians agreed on reasoning at least as well as correctness: five-rater ICC(A,k) for diagnostic reasoning was 0.773 (95% CI, 0.702 to 0.831), against a mean pairwise κ of 0.626 for correctness, although single-rater agreement was 0.41 (**Section S3**). The two measures were related within vignettes. Of 486 ratings judging a model correct, 18 (3.7%) scored its reasoning 1 or below; of 114 judging it incorrect, 34 (29.8%) scored its reasoning 3 or above, against 82.1% of correct ratings. The divergence lies between models rather than within vignettes. One clinician’s comments identified unsafe, stigmatizing, or fabricated content in four traces, all from Gemini 3 Pro (**Section S6**); in three of the four traces, all five raters judged the diagnosis correct. Automated accuracy was fragile

Three features of the pipeline moved automated accuracy by amounts comparable to the differences between models. First, the adjudicator agreed with the clinician majority on 82.5% of the 120 pairs (95% CI, 73.3 to 90.0; κ = 0.52), less closely than individual clinicians agreed with held-out majorities (0.89 to 0.96, against 0.83 to 0.85 for the adjudicator). It rejected 16 diagnoses the majority accepted, most often over a specifier or code, and accepted 5 it rejected (*p* = 0.027), lowering estimated accuracy by up to 17 percentage points depending on the model (**Section S4**). Second, outputs and verdicts were unstable. Two generation runs four days apart changed the top-1 verdict for 18 of 119 pairs (15.1%) and which model ranked best, and three adjudications of identical outputs moved accuracy by up to 5.6 points and changed how many pairs were separated on top-5 accuracy (zero, two, and one). Only Claude Opus 4.5 was accessed through a dated snapshot, so for the other models run-to-run variation cannot be separated from provider updates. Third, scoring depended on how references were written. Nine reference strings listed several diagnoses in one string, which no single predicted diagnosis could match; splitting them raised each model’s top-5 accuracy by 2.2 to 2.6 points and changed one pairwise result (**Section S4**).

### Trace features did not substitute for clinician ratings

Across the four model means, the number of distinct ICD-10 codes named in a trace was strongly inversely associated with reasoning ratings (ρ = -0.80), although with four models no association can reach a two-sided *p* below 0.083. Across the 120 traces, with a random intercept for each vignette, the features were null, reversed, or too small to explain the differences (**Figure 3**). Each additional code was associated with +0.02 points (95% CI, -0.01 to +0.05), reversing the model-level sign; and trace length predicted at most 0.27 points of the observed 1.23 range. The share of traces naming no code ranged from 0% to 53% by model, and the lowest-rated system named codes in every trace: the aggregate feature tracked formatting, not reasoning (**Table S5C**).

**Figure 3.**
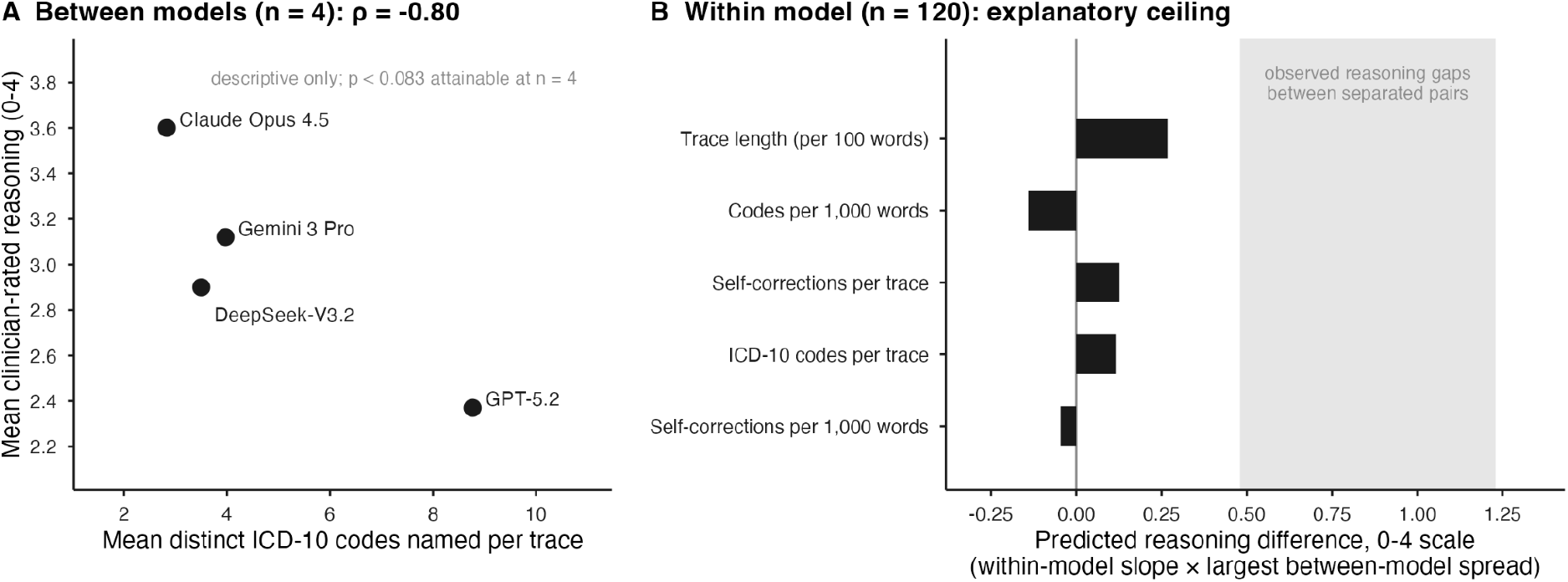
Computable features do not substitute for clinician assessment of reasoning traces (*n* = 30) **(A)** Across the four model means, the number of distinct ICD-10 codes named per trace was inversely associated with clinician-rated reasoning (Spearman ρ = −0.80); with four models, no exact two-sided p below 0.083 is attainable, so the association is descriptive. **(B)** For each feature, the within-model coefficient from a linear mixed model (reasoning ∼ feature + model, random intercept for vignette, 120 traces) multiplied by the largest between-model spread in that feature, which gives the largest reasoning difference the feature could explain. The shaded band spans the observed reasoning differences between separated pairs (0.48 to 1.23 points). Coefficients and confidence intervals are in **Table S5C**.

## Discussion

Accuracy did not tell these models apart, and clinician-rated reasoning did. Two systems that were correct equally often, within a bounded margin, differed by 0.70 points on a 0-4 scale, and every rater placed them in the same order. For a classification tool this may not matter. For a cognitive aid, where a clinician decides whether to accept or question an output, two equally accurate models may not be clinically equivalent. We have not shown an effect on clinician decisions,^39^ which requires prospective study.

Automated accuracy was a moving target. Re-running the adjudicator alone moved accuracy by up to 5.6 points, more than the differences among the three lower-ranked models (0.8 to 2.5 points on top-5), and regeneration and reference formatting moved it further; the adjudicator also under-credited three of four models relative to clinicians. Evaluations built on served APIs, which change without notice, should report this variation alongside their estimates and audit how references are written before scoring.^16–18,20,38^

Clinician ratings could not be replaced by computable features. A feature that ranked the models as clinicians did across model averages was null within models, because it measured whether a model formats its differential with codes. This ecological fallacy is likely to recur wherever aggregate trace features are proposed as substitutes for expert judgment, and it argues for validating any such proxy at the level at which it will be used. Panel rating was itself tractable: five-rater agreement on reasoning was at least as high as agreement on diagnostic correctness and rater variance was negligible, although single raters were not reliable enough to replace a panel.

This evaluation has limitations. First, correctness and reasoning were rated within the same sessions, so knowledge of correctness may have influenced reasoning scores; a halo would pull equally accurate models together, biasing against our result, but does not substitute for rating reasoning before correctness is known. Second, the reasoning findings rest on 30 vignettes, one of them written by a rater, and equivalence was established for one pair only, which automated top-1 accuracy on the full set favored by 9.5 points, significantly in one of three passes. Third, the reference standard is imperfect: vignettes were screened by single reviewers, some reference diagnoses list alternatives, and an independent psychiatrist disputed one (bipolar I rather than II disorder). Published case reports overrepresent unusual presentations, which limits inference about routine practice but samples the diagnostic long tail densely, and similar accuracy across sources does not exclude memorization, since the sources differ in case mix and length. Fourth, static vignettes isolate reasoning from history-taking, and we rated single, uninterrogated responses rather than the iterative use clinicians are advised to adopt,^20^ so whether the differences persist when models must gather information requires interactive evaluation.^40^ Fifth, we rated provider-supplied trace summaries elicited under identical explicit guidance, not internal computation or unprompted reasoning, although these summaries are what a clinician would audit.

## Conclusions

Four LLMs that accuracy separated weakly or not at all differed consistently in clinician-rated reasoning, and computable trace features did not recover the difference. Where models are meant to support clinical reasoning, pre-deployment evaluation should include panel-based clinician review of reasoning traces and report how stable its accuracy estimates are.

## Data availability

Analysis code, evaluation prompts, and derived evaluation outputs will be [publicly available in a repository](https://github.com/BIDS-Xu-Lab/psychiatry-frontier-llm-evaluation) upon publication. Clinician-authored fictitious vignettes will also be publicly available. We will not publicly redistribute text derived from published case reports or verbatim model reasoning traces; citations to original sources of the case reports will be provided, and access to restricted materials may be provided under controlled conditions (e.g., to qualified researchers under a data use agreement and/or institutional approval).

## Acknowledgments

The authors thank Daniela Weng for creating Figure 1, and Qianqian Xie, Lingfei Qian, and Jeffrey Zhang for their advice on study design. This study received material support in the form of API credits from the OpenAI Researcher Access Program and the Google Gemini Academic Program. KWJ is supported by the National Science Foundation’s Graduate Research Fellowship Program and was formerly supported by the National Library of Medicine’s T15 University-based Biomedical Informatics and Data Science Training Program for a portion of this study. MMal was supported by the National Institute of Mental Health under grant K23MH134068. JK is supported by the National Institute of Mental Health under grant K01MH137386. The content is solely the responsibility of the authors and does not necessarily represent the official views of the National Institutes of Health. The funders had no role in study design, model selection, data collection, data analysis, data interpretation, or writing of the manuscript. In the last 3 years, CIR has served as a consultant for Biohaven Pharmaceuticals and Osmind; and receives research grant support from Biohaven Pharmaceuticals, a stipend from American Psychiatric Association Publishing for her role as Deputy Editor at The American Journal of Psychiatry, a stipend from Springer Nature for her role as Deputy Editor for Neuropsychopharmacology, and book royalties from American Psychiatric Association Publishing. The remaining authors report no financial or other relationship relevant to the subject of this manuscript. Claude Opus 5.5 (Anthropic) was used to help structure and revise the manuscript and supplement, and to write analysis code; all analyses were run, and all text was checked, by the authors, who take responsibility for the content.

## Author contributions

KWJ, MSS, CP, SWY, BAZ, MMal, JK, and HX conceived and designed the study. KWJ and HX acquired funding. KWJ was responsible for creating all figures and tables, except for Figure 1. CN, JW, JSZ, MAG, ASH, MMon, and JS curated data. JW, CN, ASH, JSZ, PC, and BAZ wrote fictitious vignettes. CIR, SS, WSM, CN, and PC evaluated clinical reasoning. JSZ and YRA served as the trainee comparison. MSS rated diagnostic ambiguity of the 30-case vignette subset. KWJ acquired, analyzed, and interpreted the data. KWJ and QC have directly accessed and verified the underlying data reported in the manuscript. All authors contributed to drafting or critical review of the manuscript, approved the final version, and accept responsibility for the decision to submit for publication.

