## Supplementary Appendix for "Diagnostic accuracy and clinician-rated reasoning quality dissociate in a blinded evaluation of large language models in psychiatry"

Regeneration 18
**Section S5: Statistical results 19**
Methods 19
Discordant cases 19
Table S5A: Pairwise comparisons of clinician-determined accuracy and clinician-rated reasoning (n = 30 vignettes) 19
Table S5B: Sensitivity analyses of the pairwise comparisons 20
Table S5C: Computable trace features and clinician-rated reasoning 20
**Section S6: Qualitative analysis 22**
Pipeline 22
Table S6A: Domain-level clinician concerns with model reasoning out of all comments 22
Table S6B: Rule-based detailed model reasoning failure modes out of all comments 22
Table S6C: Reasoning theme prevalence by model out of non-boilerplate comments 23
Unsafe, stigmatizing, or fabricated content 24
**Section S7: Trainee comparison 25**
Table S7: Comparison of diagnostic accuracy between Claude Opus 4.5 and two psychiatry residents on the 30 rated vignettes 25

### Section S1: Dataset

<u>Published vignette curation criteria</u>

The set of case reports derived from medical literature was randomly split into seven sections. Each section was then curated by a different clinician using inclusion criteria determined through clinician consensus (reproduced below). Each clinician rated cases in their section on a 1 (worst) to 5 (best) Likert scale, retaining cases that scored 3 or higher.

The seven screeners were two board-certified psychiatrists and five psychiatry residents. Each assigned one composite score per case for diagnostic clarity, narrative coherence, and completeness of the information required to reach the reference diagnosis, rather than scoring these dimensions separately. The 48 of 183 cases (26.2%) scoring below 3 were excluded, leaving 135, and study authors then removed explicit diagnostic labels from the retained cases.

5 = Highly appropriate
- Clear presentation of symptoms and relevant history
- Realistic case that could be encountered in practice
- Contains sufficient information for diagnosis
- May be challenging but in a clinically relevant way

4 = Moderately appropriate
- Generally good case with minor issues
- Might benefit from small clarifications
- Still represents a fair test of diagnostic ability

3 = Neutral
- Acceptable but has notable issues
- May need moderate revision to be ideal
- Still potentially usable with modifications

2 = Somewhat problematic
- Significant issues with clarity or realism
- Missing important clinical information
- May not represent a fair test of diagnostic ability

1 = Not appropriate
- Unclear or confusing presentation
- Highly unrealistic scenario
- Missing crucial diagnostic information
- Would not fairly test diagnostic capability

Per-vignette quality scores for published vignettes

| Vignette | Rating |
|---|---|
| 1 | 2 |
| 2 | 5 |
| 3 | 1 |
| 4 | 5 |
| 5 | 2 |
| 6 | 2 |
| 7 | 2 |
| 8 | 1 |
| 9 | 0 |
| 10 | 2 |
| 11 | 2 |
| 12 | 1 |
| 13 | 4 |
| 14 | 3 |
| 15 | 3 |
| 16 | 1 |
| 17 | 4 |
| 18 | 4 |
| 19 | 5 |
| 20 | 3 |
| 21 | 5 |
| 22 | 5 |
| 23 | 5 |
| 24 | 4 |
| 25 | 4 |
| 26 | 5 |
| 27 | 5 |
| 28 | 4 |
| 29 | 0 |
| 30 | 5 |
| 31 | 1 |
| 32 | 3 |
| 33 | 3 |
| 34 | 4 |
| 35 | 3 |
| 36 | 4 |
| 37 | 5 |
| 38 | 1 |
| 39 | 5 |
| 40 | 5 |
| 41 | 2 |
| 42 | 2 |
| 43 | 5 |
| 44 | 5 |
| 45 | 2 |
| 46 | 1 |
| 47 | 2 |
| 48 | 5 |
| 49 | 2 |
| 50 | 5 |
| 51 | 1 |
| 52 | 1 |
| 53 | 1 |
| 54 | 2 |
| 55 | 1 |
| 56 | 2 |
| 57 | 2 |
| 58 | 2 |
| 59 | 2 |
| 60 | 4 |
| 61 | 2 |
| 62 | 5 |
| 63 | 1 |
| 64 | 5 |
| 65 | 2 |
| 66 | 4 |
| 67 | 1 |
| 68 | 0 |
| 69 | 2 |
| 70 | 3 |
| 71 | 2 |
| 72 | 4 |
| 73 | 4 |
| 74 | 2 |
| 75 | 2 |
| 76 | 2 |
| 77 | 5 |
| 78 | 3 |
| 79 | 2 |
| 80 | 5 |
| 81 | 5 |
| 82 | 4 |
| 83 | 3 |
| 84 | 2 |
| 85 | 5 |
| 86 | 5 |
| 87 | 5 |
| 88 | 2 |
| 89 | 5 |
| 90 | 5 |
| 91 | 4 |
| 92 | 4 |
| 93 | 4 |
| 94 | 4 |
| 95 | 5 |
| 96 | 4 |
| 97 | 4 |
| 98 | 2 |
| 99 | 5 |
| 100 | 4 |
| 101 | 4 |
| 102 | 5 |
| 103 | 5 |
| 104 | 5 |
| 105 | 5 |
| 106 | 5 |
| 107 | 4 |
| 108 | 5 |
| 109 | 5 |
| 110 | 5 |
| 111 | 4 |
| 112 | 2 |
| 113 | 5 |
| 114 | 5 |
| 115 | 3 |
| 116 | 4 |
| 117 | 4 |

| | | | | | |
|---|---|---|---|---|---|
| 118 | 3 | 140 | 2 | 162 | 5 |
| 119 | 5 | 141 | 3 | 163 | 3 |
| 120 | 4 | 142 | 5 | 164 | 1 |
| 121 | 5 | 143 | 2 | 165 | 5 |
| 122 | 5 | 144 | 4 | 166 | 5 |
| 123 | 5 | 145 | 5 | 167 | 4 |
| 124 | 3 | 146 | 3 | 168 | 5 |
| 125 | 5 | 147 | 4 | 169 | 5 |
| 126 | 5 | 148 | 5 | 170 | 5 |
| 127 | 5 | 149 | 4 | 171 | 4 |
| 128 | 3 | 150 | 5 | 172 | 5 |
| 129 | 5 | 151 | 3 | 173 | 4 |
| 130 | 3 | 152 | 5 | 174 | 4 |
| 131 | 5 | 153 | 5 | 175 | 5 |
| 132 | 5 | 154 | 4 | 176 | 5 |
| 133 | 3 | 155 | 5 | 177 | 5 |
| 134 | 3 | 156 | 5 | 178 | 5 |
| 135 | 4 | 157 | 5 | 179 | 4 |
| 136 | 5 | 158 | 4 | 180 | 5 |
| 137 | 4 | 159 | 5 | 181 | 5 |
| 138 | 4 | 160 | 4 | 182 | 5 |
| 139 | 4 | 161 | 4 | 183 | 1 |

Three cases (vignettes 9, 29, and 68) were scored 0 by one screener, below the scale minimum; they were treated as 1 and excluded with the other cases scoring below 3.

Limitations of published vignette screening procedure
A single reviewer screened each vignette, so interrater agreement on screening cannot be estimated. Also, reviewers rendered a composite judgment rather than scoring face validity, content validity, and internal consistency as separate dimensions; the criteria span those constructs but do not measure them independently.

Authoring
Clinicians were encouraged to base their vignettes almost entirely on their lived clinical experience, aiming to write vignettes that approximated real cases, with identifying information (i.e., names, dates) replaced. They were informed that the vignettes would merge into a larger dataset for future public release. They were instructed to reference the following guidelines:

Guidelines for creating new vignettes:

- Length: Similar to existing dataset (approximately 1-3 paragraphs)
- Content: Include key clinical features, relevant history, and presenting symptoms
- Diagnosis: Use standardized DSM diagnoses with ICD-10 F-codes
- Diversity: Mix of common and fringe cases from your clinical experience
- Clinical Reasoning: Include your diagnostic reasoning process (optional but valuable)

Additional considerations:

- Include both straightforward and complex cases
- Ensure patient privacy by creating fully fictitious cases
- Draw from your clinical experience while maintaining realism
- Consider including cases that might challenge LLM diagnostic capabilities

Each author wrote approximately 10 vignettes, ranging from 6 to 15.

The six authors were doctoral-level clinicians: two board-certified psychiatrists, three psychiatry residents, and one board-certified clinical psychologist. One author (PC) was also a reasoning rater, and one of these vignettes (1056) was among the 30 rated in the rated subset.

Vignette source comparison
Top-1 and top-5 accuracy were compared between published and clinician-authored vignettes using the majority verdict of the three adjudication passes at the fuzzy threshold of 90 (see **Section S4** for explanation). Per model, sources were compared with Fisher's exact test. Across models, correctness was modeled in a single mixed-effects logistic regression with fixed effects for model, vignette source, and their interaction and a random intercept for vignette, fitted by maximum likelihood with adaptive Gauss-Hermite quadrature (lme4::glmer, 25 points); likelihood-ratio tests assessed whether the source effect differed by model (3 df) and whether there was a source effect averaged over models (1 df). A population-averaged model (generalized estimating equations, exchangeable correlation within vignette) was fitted as a check. Odds ratios greater than 1 indicate higher accuracy on published vignettes, the direction expected if models had memorized published cases.

**Figure S1: Diagnostic accuracy of large language models on clinician-authored versus published vignettes**

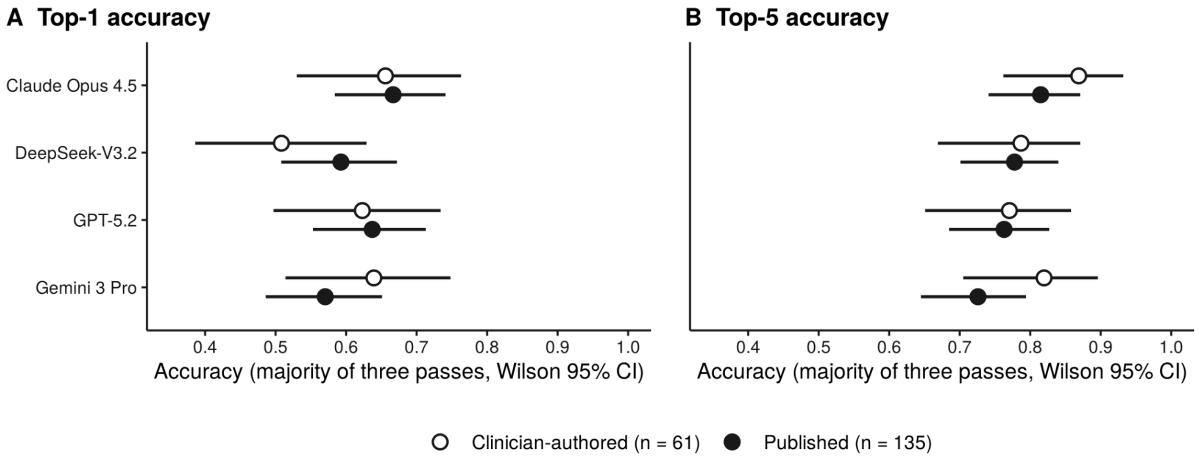

Majority verdict of three adjudication passes at the fuzzy threshold of 90, with reference strings combining several diagnoses split before scoring; bars are Wilson 95% confidence intervals. Counts and Fisher's exact tests are in **Table S1A**, and the mixed-effects model in **Table S1B**.

**Table S1A: Diagnostic accuracy by vignette source (n = 196; majority verdict of three adjudication passes, Wilson 95% CI)**

| **Model** | **Source** | **Top-1 correct** | **Top-1 accuracy (95% CI)** | **Top-5 correct** | **Top-5 accuracy (95% CI)** |
|---|---|---|---|---|---|
| Claude Opus 4.5 | Clinician-authored (n = 61) | 40 | 0.656 (0.530-0.763) | 53 | 0.869 (0.762-0.932) |
| | Published (n = 135) | 90 | 0.667 (0.584-0.741) | 110 | 0.815 (0.741-0.871) |
| DeepSeek-V 3.2 | Clinician-authored | 31 | 0.508 (0.386-0.629) | 48 | 0.787 (0.669-0.871) |
| | Published | 80 | 0.593 (0.508-0.672) | 105 | 0.778 (0.701-0.840) |
| GPT-5.2 | Clinician-authored | 38 | 0.623 (0.497-0.734) | 47 | 0.770 (0.651-0.858) |
| | Published | 86 | 0.637 (0.553-0.713) | 103 | 0.763 (0.685-0.827) |
| Gemini 3 Pro | Clinician-authored | 39 | 0.639 (0.514-0.748) | 50 | 0.820 (0.705-0.896) |
| | Published | 77 | 0.570 (0.486-0.651) | 98 | 0.726 (0.645-0.794) |

*Fisher's exact p, published versus clinician-authored, top-1: Claude Opus 4.5 0.87, DeepSeek-V3.2 0.28, GPT-5.2 0.87, Gemini 3 Pro 0.43; top-5: 0.41, 1.00, 1.00, 0.21. Unadjusted. Reference strings combining several diagnoses were split before scoring (**Section S4**).*

**Table S1B: Mixed-effects logistic regression of diagnostic correctness on vignette source and model (n = 196 vignettes × 4 models)**

| **Estimate** | **Top-1 odds ratio (95% CI)** | ***p*** | **Top-5 odds ratio (95% CI)** | ***p*** |
|---|---|---|---|---|
| Source effect, all models (no interaction) | 1.18 (0.49–2.80) | 0.71 | 0.74 (0.23–2.41) | 0.61 |

| | | | | |
|---|---|---|---|---|
| Claude Opus 4.5 | 1.22 (0.39–3.87) | 0.73 | 0.60 (0.12–3.04) | 0.54 |
| DeepSeek-V3.2 | 1.96 (0.64–6.02) | 0.24 | 1.08 (0.24–4.76) | 0.92 |
| GPT-5.2 | 1.23 (0.39–3.84) | 0.72 | 1.07 (0.25–4.64) | 0.92 |
| Gemini 3 Pro | 0.64 (0.21–1.99) | 0.44 | 0.40 (0.09–1.78) | 0.23 |
| Population-averaged check (GEE), all models | 1.05 (0.66–1.66) | 0.85 | 0.78 (0.45–1.38) | 0.40 |
| Source × model interaction, likelihood-ratio test (3 df) | | 0.31 | | 0.47 |

*Odds ratios compare published with clinician-authored vignettes; per-model rows are conditional (vignette-specific) estimates from the interaction model, with Wald intervals. Vignette random-intercept SD: 2.42 (top-1), 3.10 (top-5). Per pass, the top-1 interaction p was 0.86, 0.12, and 0.33, and the pooled top-1 odds ratio 1.40, 1.28, and 1.34. A fit integrating the random intercept numerically in Python gave the same estimates to within 0.2%. The two sources differ in case mix and length (**Tables S1D** and **S1E**), so similar accuracy does not by itself exclude memorization.*

Ambiguity

A psychiatrist who did not rate reasoning traces (MSS) scored each of the 30 vignettes rated in the rated subset for diagnostic ambiguity, from 0 (a single clear diagnosis) to 4 (no diagnosis confidently establishable), and named any competing diagnosis. Twenty-three vignettes scored 0, one scored 1, two scored 2, three scored 3, and one scored 4. Six of the seven vignettes scored above 0 were published case reports (6 of 19), against 1 of 11 clinician-authored vignettes. Because most vignettes scored 0, ambiguity was analyzed as absent or present. For one vignette scored 0 (vignette 94), the rater noted that the reference diagnosis was incorrect rather than ambiguous; this vignette is examined separately in **Table S5B**.

Ambiguity was rated only for the 30 vignettes rated in the rated subset, because its purpose was to interpret clinician disagreement and reasoning scores on those vignettes. It was rated by a psychiatrist who did not rate reasoning, so that ambiguity and reasoning ratings could not reflect the same clinician's judgment.

**Table S1C: Diagnostic ambiguity of the 30 clinician-rated vignettes**

| Vignette | Source | Ambiguity (0–4) | Competing diagnosis | Rater note |
|---|---|---|---|---|
| 17 | Published | 3 | Bipolar Disorder | |
| 23 | Published | 0 | | |
| 32 | Published | 4 | Borderline Personality Disorder, Traumatic Brain Injury | |
| 62 | Published | 3 | Seizure Disorder | |
| 80 | Published | 0 | | |
| 85 | Published | 0 | | |

| | | | | |
|---|---|---|---|---|
| 94 | Published | 0 | | The primary diagnosis listed here is incorrect. The patient clearly describes history of manic episodes and meets criteria for Bipolar 1 disorder, most recent episode depressed. |
| 97 | Published | 3 | Bipolar 1 Disorder | |
| 99 | Published | 0 | | |
| 104 | Published | 0 | | |
| 108 | Published | 2 | Major Depressive Disorder | |
| 109 | Published | 0 | | |
| 115 | Published | 0 | | |
| 118 | Published | 0 | | |
| 155 | Published | 0 | | |
| 159 | Published | 0 | | |
| 169 | Published | 0 | | |
| 178 | Published | 2 | | Not enough information to make any diagnosis apart from Benzodiazepine dependence |
| 181 | Published | 0 | | |
| 1003 | Clinician-authored | 0 | | |
| 1009 | Clinician-authored | 0 | | |
| 1012 | Clinician-authored | 0 | | |
| 1013 | Clinician-authored | 0 | | |
| 1020 | Clinician-authored | 0 | | |
| 1021 | Clinician-authored | 0 | | |
| 1032 | Clinician-authored | 0 | | |

| | | | | |
|---|---|---|---|---|
| 1033 | Clinician-authored | 1 | | Not enough information for a clear diagnosis |
| 1047 | Clinician-authored | 0 | | |
| 1049 | Clinician-authored | 0 | | |
| 1056 | Clinician-authored | 0 | | |

*Scores from one rater.*

<u>Case mix</u>

Each vignette's primary reference diagnosis (the first listed) was assigned to its DSM-5-TR diagnostic class; vignettes whose primary reference was a medical condition presenting with psychiatric symptoms were classed separately. Primary references spanned 19 DSM-5-TR diagnostic classes, and all reference diagnoses together (281 across 196 vignettes) spanned the same classes plus one condition for further study. Sixty-two vignettes (32%) listed more than one reference diagnosis, and in 15 (8%) the primary reference was a medical condition. The four most common classes (schizophrenia spectrum, depressive, substance-related, and bipolar disorders) accounted for 80 vignettes (41%). Clinician-authored vignettes were weighted towards obsessive-compulsive disorders (10 of 61, against 5 of 135 published) and contained no somatic symptom, dissociative, paraphilic, sexual dysfunction, sleep-wake, gender dysphoria, or elimination disorders as the primary reference. The 30 vignettes rated in the rated subset covered 14 of the 21 categories; the largest class absent from them was trauma- and stressor-related disorders (16 vignettes in the full set).

**Table S1D: Primary reference diagnosis by DSM-5-TR diagnostic class and vignette source**

| **Primary reference diagnosis, DSM-5-TR class** | **Clinician-authored (n = 61)** | **Published (n = 135)** | **All (n = 196)** | **Rated subset (n = 30)** |
|---|---|---|---|---|
| Schizophrenia spectrum and other psychotic | 9 | 15 | 24 (12.2%) | 4 |
| Depressive | 8 | 15 | 23 (11.7%) | 4 |
| Substance-related and addictive | 6 | 11 | 17 (8.7%) | 4 |
| Bipolar and related | 6 | 10 | 16 (8.2%) | 4 |
| Trauma- and stressor-related | 6 | 10 | 16 (8.2%) | 0 |
| Obsessive-compulsive and related | 10 | 5 | 15 (7.7%) | 2 |
| Personality | 3 | 10 | 13 (6.6%) | 1 |
| Somatic symptom and related | 0 | 9 | 9 (4.6%) | 1 |
| Anxiety | 1 | 7 | 8 (4.1%) | 1 |
| Neurodevelopmental | 2 | 6 | 8 (4.1%) | 3 |
| Neurocognitive | 1 | 6 | 7 (3.6%) | 1 |
| Feeding and eating | 2 | 5 | 7 (3.6%) | 1 |

| | | | | |
|---|---|---|---|---|
| Disruptive, impulse-control, and conduct | 4 | 2 | 6 (3.1%) | 2 |
| Dissociative | 0 | 3 | 3 (1.5%) | 0 |
| Paraphilic | 0 | 2 | 2 (1.0%) | 1 |
| Sexual dysfunctions | 0 | 2 | 2 (1.0%) | 0 |
| Sleep-wake | 0 | 2 | 2 (1.0%) | 0 |
| Gender dysphoria | 0 | 1 | 1 (0.5%) | 0 |
| Elimination | 0 | 1 | 1 (0.5%) | 0 |
| Medical condition (non-psychiatric) | 3 | 12 | 15 (7.7%) | 1 |
| No diagnosis | 0 | 1 | 1 (0.5%) | 0 |

Reference diagnoses

Reference strings were reviewed for features that affect scoring. In 7 vignettes the reference named alternatives ("or", "vs"); in 17, a single reference string named diagnoses from two or more classes and was therefore scored as one reference; in 15, the primary reference was not a mental disorder; in 4, a reference was provisional, a rule-out, or historical; and in 2, the reference text contained a parsing anomaly. Because a vignette can carry more than one of these features, the 45 instances affected 37 vignettes (19%). Reference strings combining several diagnoses were split before scoring (**Section S4**).

Sixty-two vignettes (32%) listed more than one reference diagnosis; a prediction was scored correct if it matched any of them, and recall@5 counts the proportion recovered.

Length and temporal course

Vignettes had a median length of 453 words (IQR, 321 to 576; range, 143 to 1,290). Clinician-authored vignettes were shorter than published ones (median 314 words, IQR 239 to 348, against 533, IQR 421 to 624). The 30 rated vignettes were similar in length to the 166 that were not rated (median 447 and 453 words).

Vignettes were presented to models as written. Diagnoses whose criteria require a minimum duration or a particular course therefore depended on the history each vignette gave; no temporal information was added, and temporal course was not coded separately.

**Table S1E: Vignette length in words**

| | **n** | **Median** | **IQR** | **Range** |
|---|---|---|---|---|
| All vignettes | 196 | 453 | 321–576 | 143–1,290 |
| Clinician-authored | 61 | 314 | 239–348 | 143–598 |
| Published | 135 | 533 | 421–624 | 172–1,290 |
| Rated in the rated subset | 30 | 447 | 353–585 | 171–1,138 |
| Not rated | 166 | 453 | 318–575 | 143–1,290 |

**Section S2: Models and prompts**

Prompts

**System instructions**

You are an expert psychiatrist with extensive clinical experience. Your responsibility is to accurately diagnose patients based on clinical case vignettes while following diagnostic criteria established in the Diagnostic and Statistical Manual of Mental Disorders, Fifth Edition, Text Revision (DSM-5-TR). Allow for diagnostic flexibility in rare or complex cases.

**User prompt**

As you are reading the clinical case vignette and building your differential diagnosis, follow the below clinical reasoning rubric adapted from the ACGME Psychiatry Milestones, which evaluates psychiatry resident physician performance. The rubric is enclosed between <rubric> and </rubric>. This rubric contains two subcompetencies - Data Extraction and Organization, and Diagnostic Reasoning and Differential Diagnosis. You should perform at Level 5 for both, representing an expert resident whose achievements in a subcompetency are greater than the expectation.

<rubric>

A - Data Extraction and Organization: Does the diagnostician effectively extract relevant information from the case vignette and organize it logically to support their reasoning?

1: Fails to identify key clinical details; the extracted information is inaccurate, irrelevant, or nonsensical.

2: Identifies some basic symptoms but misses crucial contextual information (timeline, triggers, associated features, risk factors). Organization is poor or absent.

3: Extracts most of the core symptoms and some contextual details. Presents the information in a basic structured format (eg, lists symptoms, some history).

4: Accurately extracts and organizes the key clinical information, including relevant history, mental status findings, and risk factors, in a clear and logical structure.

5: Demonstrates a sophisticated ability to identify subtle cues, patterns, and relationships within the clinical data and organizes them in a way that clearly supports a nuanced understanding of the case.

B - Diagnostic Reasoning and Differential Diagnosis: Does the diagnostician generate a plausible differential diagnosis and provide a logical rationale connecting the clinical information to its diagnostic considerations?

1: Provides no diagnosis or a diagnosis that is completely unrelated to the clinical information presented. The reasoning is absent or entirely illogical.

2: States a diagnosis with minimal or no justification, or the justification provided is weak, tautological, or contradicts the clinical data. Fails to consider alternative diagnoses.

3: Generates a basic differential diagnosis that includes the most likely condition, with some attempt to link symptoms to diagnostic criteria. The reasoning is present but may be superficial or incomplete.

4: Develops a reasonable differential diagnosis, prioritizing conditions based on the presented information. Provides a clear rationale, explicitly linking key clinical features to each diagnostic consideration. May briefly mention rule-outs.

5: Constructs a sophisticated and nuanced differential diagnosis, considering less obvious possibilities and providing a well-articulated rationale that demonstrates a deep understanding of psychopathology and diagnostic principles. Clearly articulates the reasoning for prioritizing certain diagnoses and ruling out others.

</rubric>

Based on the information above, please read and diagnose the following clinical case vignette enclosed between the <vignette> and </vignette> tags, and provide a structured list of DSM-5-TR psychiatric diagnoses for it. Base your answer only on the information provided and your clinical expertise; do not fabricate additional details. Give the top 5 most likely diagnoses with corresponding ICD-10 F-codes, consisting of the primary diagnosis and 4 secondary diagnoses with corresponding ICD-10 F-codes. Do not include non-psychiatric diagnoses that do not use F-codes. Do not output any of your clinical reasoning; your response should consist only of the structured list of diagnoses and corresponding F-codes. Follow this formatting:

1. <Primary diagnosis> - ICD-10 F-code
2. <Secondary diagnosis> - ICD-10 F-code
3. <Secondary diagnosis> - ICD-10 F-code

**Section S3: Clinician rating**

Correctness instruction supplied to clinicians

Verify if the diagnostician's predicted diagnosis matches the true diagnosis (Yes/No). Answer Yes if the model has captured at least the primary diagnosis in its response.

Reasoning rubric

Note: While the prompt provided to the LLMs utilized the standard ACGME 1-5 scale to induce "expert resident behavior," human evaluators utilized a 0-4 scale. This allowed the expert judges to assign a 0 for completely hallucinatory or nonsensical outputs, which shifted the standard ACGME 1-5 scale down by one point.

**Data Extraction and Organization**

Does the diagnostician effectively extract relevant information from the case vignette and organize it logically to support their reasoning?

- **1:** Fails to identify key clinical details; the extracted information is inaccurate, irrelevant, or nonsensical.
- **2:** Identifies some basic symptoms but misses crucial contextual information (timeline, triggers, associated features, risk factors). Organization is poor or absent.
- **3:** Extracts most of the core symptoms and some contextual details. Presents the information in a basic structured format (e.g., lists symptoms, some history).
- **4:** Accurately extracts and organizes the key clinical information, including relevant history, mental status findings, and risk factors, in a clear and logical structure.
- **5:** Demonstrates a sophisticated ability to identify subtle cues, patterns, and relationships within the clinical data and organizes them in a way that clearly supports a nuanced understanding of the case.

**Diagnostic Reasoning and Differential Diagnosis**

Does the diagnostician generate a plausible differential diagnosis and provide a logical rationale connecting the clinical information to its diagnostic considerations?

- **1:** Provides no diagnosis or a diagnosis that is completely unrelated to the clinical information presented. The reasoning is absent or entirely illogical.
- **2:** States a diagnosis with minimal or no justification, or the justification provided is weak, tautological, or contradicts the clinical data. Fails to consider alternative diagnoses.
- **3:** Generates a basic differential diagnosis that includes the most likely condition, with some attempt to link symptoms to diagnostic criteria. The reasoning is present but may be superficial or incomplete.
- **4:** Develops a reasonable differential diagnosis, prioritizing conditions based on the presented information. Provides a clear rationale, explicitly linking key clinical features to each diagnostic consideration. May briefly mention rule-outs.
- **5:** Constructs a sophisticated and nuanced differential diagnosis, considering less obvious possibilities and providing a well-articulated rationale that demonstrates a deep understanding of psychopathology and diagnostic principles. Clearly articulates the reasoning for prioritizing certain diagnoses and ruling out others.

Qualitative prompts

1. Was the reasoning logically coherent?
2. Were any unsafe, stigmatizing, or hallucinated outputs present?
3. Does the diagnostician demonstrate flexibility when the data is ambiguous?

Rater background questions

1. Please describe your level of experience in psychiatry and the extent of your practice. (eg, 10+ years experience in clinical psychiatry, with specialization in OCD)
2. How often do you use LLMs?
3. What is your opinion of LLM usage in psychiatry?

Rater reliability

Interrater agreement was assessed with mean pairwise Cohen's κ for human-judged diagnostic correctness and single-rater and five-rater intraclass correlation coefficients for the human-judged data extraction score and human-judged diagnostic ability score. We report both single-rater and average-rater ICC forms to justify the use of panel-based rating: single-rater agreement was 0.41-0.51, while five-rater averaged agreement was 0.773 (95% CI 0.702-0.831) for diagnostic reasoning and 0.840 (0.790-0.881) for data extraction.

Clinicians agreed substantially on diagnostic correctness (mean pairwise κ = 0.626; range across the 10 rater pairs, 0.46 to 0.83), although 28 of 120 traces (23.3%) drew a non-unanimous verdict, with disagreement spread across all 10 rater pairs. Agreement on reasoning was therefore at least as high as agreement on correctness.

All interrater agreement metrics were implemented and calculated in R (version 4.5.2) using the "irr" package.

In a crossed random-effects model with random intercepts for vignette and rater, variance attributable to raters was 0.002, against 0.129 for vignettes and 0.625 residual: averaging across the five raters loses almost nothing, and comparing models paired within vignette removes vignette difficulty as a source of variance.

### Section S4: Adjudication

Prompt

Your task is to act as a strict medical adjudicator specializing in psychiatry and identify whether the predicted diagnosis is clinically equivalent to (or a valid subclass of) the true diagnosis. Your standards are exacting, and you must consider the nuances of each diagnosis carefully. As much as possible, adhere to the diagnostic language laid out in the DSM-5-TR, and utilize the included ICD-10 F-codes to aid your determination.

True Diagnosis: "{t}"
Predicted Diagnosis: "{p}"

Return JSON ONLY: { "match": <true/false> }

Metrics and matching

Accuracy was assessed on all 196 vignettes over each model's five ranked diagnoses. A prediction counted as correct if it matched any listed reference diagnosis: top-1 accuracy reflects whether the first-ranked prediction matched; top-5 accuracy, whether any did; recall@5, the proportion of a vignette's reference diagnoses recovered; and mean reciprocal rank, the reciprocal of the rank of the first match. Each predicted diagnosis was compared with each reference diagnosis in two tiers: a string-similarity tier (token-set ratio of at least 90, referred to "fuzzy threshold of 90") accepted near-identical strings, and all other comparisons were judged by GPT-5-mini (gpt-5-mini-2025-08-07) with the prompt above.

Repeated adjudication and failed calls

Model outputs were adjudicated three times at the fuzzy threshold of 90 and three times with the fuzzy tier disabled, so that every comparison went to GPT-5-mini. Every reference-prediction comparison was logged with the tier that decided it, the adjudicator's verdict, and any error. Failed calls were retried up to four times and, if still unsuccessful, scored as non-matches, the convention of the original pipeline; the number per vignette is recorded. In the 12 runs at threshold 90, 1 to 5 calls per run failed after retry. Most failures were refusals by the adjudicator on comparisons involving paraphilic disorders (vignettes 180 and 181); the remainder were access errors. Where a model's output contained a numbered list, unnumbered lines (preambles, blank lines, or diagnosis codes that had wrapped onto their own line) were excluded before ranks were assigned; outputs without numbering were parsed as before.

Scoring every failed call as a match rather than a non-match, the most favorable treatment, changed no model's three-pass mean top-1 or top-5 accuracy by more than 0.6 percentage points and changed no pairwise result: the Holm-adjusted p for Claude Opus 4.5 versus DeepSeek-V3.2 on top-1 accuracy rose from 0.076 to 0.105, and that for Claude Opus 4.5 versus Gemini 3 Pro on top-5 accuracy remained 0.016.

Reference strings naming several diagnoses

Nine reference strings listed two to four separate diagnoses in a single string (Section S1). The adjudicator compares each prediction with each reference and asks whether they are clinically equivalent, so a prediction naming one of the listed diagnoses was scored as a non-match, whereas the same diagnoses given as separate references would each have been matched. These strings were rewritten as numbered lists of their component diagnoses, with wording unchanged, the format already used for vignettes with several references, and the nine vignettes were re-adjudicated for all four models with the same settings (three passes at each threshold). The split references are deposited with the code. All results in the manuscript and supplement use the split references. Before the split, three-pass mean top-5 accuracy was 0.811, 0.762, 0.741, and 0.733 (Claude Opus 4.5, DeepSeek-V3.2, GPT-5.2, Gemini 3 Pro) and top-1 accuracy 0.650, 0.554, 0.604, and 0.583; the majority-of-three verdict separated Claude Opus 4.5 from DeepSeek-V3.2 on top-1 accuracy (Holm-adjusted $p = 0.040$), which it did not after the split ($p = 0.076$).

Validation against clinicians

The adjudicator was validated against the clinician majority (at least 3 of 5 raters) on the 120 vignette–model pairs rated in the rated subset, using the outputs the clinicians rated. Raters applied the correctness instruction to the model's leading diagnosis: the clinician majority credited 95% of pairs in which a reference diagnosis matched the first-listed diagnosis but 30% of pairs in which it matched only a lower-ranked one. The primary comparison therefore uses the adjudicator's top-1 verdict, a choice made after examining the ratings; the planned top-5 comparison is reported below the table. We report percent agreement, Cohen's κ, prevalence-adjusted κ, and exact McNemar tests for the direction of disagreement and, as a benchmark, the agreement of each clinician and of the adjudicator with the majority of the remaining four raters, excluding 2–2 ties. Clinician determinations are the reference standard for all analyses of the 30 rated vignettes.

**Table S4A: Clinician-determined and adjudicator-determined accuracy on the 30 rated vignettes (second generation run; threshold 90, pass 1)**

| Model | Clinician rater mean | Clinician majority | Adjudicator top-1 | Difference from majority |
|---|---|---|---|---|
| Claude Opus 4.5 | 0.827 | 0.833 | 0.700 | −13.3 |
| DeepSeek-V3.2 | 0.827 | 0.800 | 0.800 | 0.0 |
| Gemini 3 Pro | 0.820 | 0.833 | 0.667 | −16.7 |
| GPT-5.2 | 0.767 | 0.767 | 0.700 | −6.7 |

*Against the clinician majority, the adjudicator's top-1 verdict agreed on 82.5% of the 120 pairs (95% CI, 73.3 to 90.0; κ = 0.52; prevalence-adjusted κ = 0.65); it rejected 16 diagnoses the majority accepted and accepted 5 it rejected (exact McNemar p = 0.027). Its agreement with each held-out four-rater majority was 0.83 to 0.85, against 0.89 to 0.96 for individual clinicians. For the planned top-5 comparison, agreement was 74.2% (95% CI, 64.2 to 84.2; κ = 0.02), with 20 pairs accepted by the adjudicator and rejected by the majority and 11 the reverse (p = 0.15). Two vignettes in the rated subset (32 and 1047) had combined reference strings; after splitting, GPT-5.2's adjudicated accuracy rose from 0.633 to 0.700, and the number of discordant pairs changed from 17 and 4 to 16 and 5. Ten rater rows differ from the adjudicated differential only by a preamble line the parser drops (vignettes 80 and 155, Gemini 3 Pro); one rater row for vignette 104 (Gemini 3 Pro) shows a different differential from the one adjudicated.*

**Table S4B: Automated accuracy on all 196 vignettes by adjudication pass**

| Model | Fuzzy tier | Top-1, passes 1 / 2 / 3 | Top-5, passes 1 / 2 / 3 |
|---|---|---|---|
| Claude Opus 4.5 | Threshold 90 | 0.663 / 0.684 / 0.658 | 0.827 / 0.857 / 0.827 |
| | Off | 0.653 / 0.648 / 0.648 | 0.811 / 0.816 / 0.811 |
| DeepSeek-V3.2 | Threshold 90 | 0.592 / 0.561 / 0.566 | 0.791 / 0.781 / 0.781 |
| | Off | 0.531 / 0.536 / 0.520 | 0.745 / 0.735 / 0.740 |
| GPT-5.2 | Threshold 90 | 0.633 / 0.597 / 0.653 | 0.776 / 0.735 / 0.791 |
| | Off | 0.597 / 0.602 / 0.612 | 0.770 / 0.760 / 0.765 |
| Gemini 3 Pro | Threshold 90 | 0.592 / 0.612 / 0.592 | 0.760 / 0.760 / 0.755 |
| | Off | 0.571 / 0.577 / 0.592 | 0.740 / 0.735 / 0.760 |

*Table 1 in the main text reports the mean of the three threshold-90 passes. Claude Opus 4.5 ranked first on all four metrics in every threshold-90 pass. Between 13 and 18 of 196 vignettes per model received different top-5 verdicts across the three threshold-90 passes, and between 12 and 22 different top-1 verdicts.*

**Table S4C: Pairwise comparisons of automated accuracy by adjudication pass (exact McNemar tests, Holm-adjusted within metric and pass)**

| Comparison | Metric | Difference, majority | Holm *p*, pass 1 | Pass 2 | Pass 3 | Majority of 3 |
|---|---|---|---|---|---|---|
| Claude Opus 4.5 vs DeepSeek-V3.2 | Top-1 | +0.097 | 0.380 | 0.009 | 0.092 | 0.076 |
| Claude Opus 4.5 vs GPT-5.2 | Top-1 | +0.031 | 1.000 | 0.100 | 1.000 | 0.942 |
| Claude Opus 4.5 vs Gemini 3 Pro | Top-1 | +0.071 | 0.201 | 0.100 | 0.238 | 0.168 |
| DeepSeek-V3.2 vs GPT-5.2 | Top-1 | −0.066 | 1.000 | 0.900 | 0.215 | 0.495 |
| DeepSeek-V3.2 vs Gemini 3 Pro | Top-1 | −0.026 | 1.000 | 0.735 | 1.000 | 0.942 |
| GPT-5.2 vs Gemini 3 Pro | Top-1 | +0.041 | 1.000 | 0.900 | 0.379 | 0.937 |
| Claude Opus 4.5 vs DeepSeek-V3.2 | Top-5 | +0.051 | 1.000 | 0.054 | 0.814 | 0.441 |
| Claude Opus 4.5 vs GPT-5.2 | Top-5 | +0.066 | 0.378 | <0.001 | 0.918 | 0.096 |
| Claude Opus 4.5 vs Gemini 3 Pro | Top-5 | +0.077 | 0.064 | <0.001 | 0.040 | 0.016 |
| DeepSeek-V3.2 vs GPT-5.2 | Top-5 | +0.015 | 1.000 | 0.698 | 1.000 | 1.000 |
| DeepSeek-V3.2 vs Gemini 3 Pro | Top-5 | +0.026 | 1.000 | 1.000 | 1.000 | 1.000 |
| GPT-5.2 vs Gemini 3 Pro | Top-5 | +0.010 | 1.000 | 1.000 | 1.000 | 1.000 |

*Threshold 90, split references. The majority-of-three test was specified after the passes were run. Pairs separated (Holm-adjusted $p < 0.05$): pass 1, none; pass 2, one on top-1 and two on top-5; pass 3, one on top-5; majority, one on top-5 and none on top-1. Differences are majority-verdict accuracies, which can differ slightly from the three-pass means in Table 1.*

Regeneration

The outputs the clinicians rated came from a second generation run made within four days of the run used for Table 1, with identical prompts, parameters, and model aliases, which permitted an unplanned test–retest comparison on 119 vignette-model pairs (one GPT-5.2 output in the first run, for vignette 94, was withheld by a provider content filter). Both runs were adjudicated with the logged pipeline (three passes at the fuzzy threshold of 90, split reference strings), and each pair's verdict is the majority of the three passes. Regeneration changed the top-1 verdict for 18 of 119 pairs (15.1%) and the top-5 verdict for 17 (14.3%); across the nine pairings of individual passes, top-1 changes ranged from 16 to 21. GPT-5.2's top-5 accuracy on these vignettes moved 10.3 percentage points between runs, the best-performing model differed between runs (Claude Opus 4.5 on both metrics in the first; DeepSeek-V3.2 on top-1 and GPT-5.2 on top-5 in the second), and the between-model spread in top-5 accuracy was 20.0 points in the first run and 16.6 in the second. Only Claude Opus 4.5 was accessed through a dated snapshot, so for the other models run-to-run variation cannot be separated from provider-side updates.

**Section S5: Statistical results**

Methods

Each vignette contributes one score per model, averaged over raters, and models were compared pairwise within vignette with 95% paired t-intervals (df = 29) and Holm-Bonferroni adjustment across the six comparisons per measure; estimates were cross-checked against a crossed random-effects model with random intercepts for vignette and rater. Because failure to reject a difference is not evidence of equivalence, accuracy equivalence was tested by two one-sided tests, reporting for each pair the smallest margin at which equivalence holds, unadjusted and Holm-adjusted, with bootstrap percentile intervals as a check; the pair carrying the primary claim was selected because its accuracies matched, which is post hoc. Full-set automated accuracy was compared between models with exact McNemar tests on the majority verdict of the three adjudication passes, Holm-adjusted within metric, a test specified after the passes were run; Table 1 intervals are bootstrap percentile intervals over vignettes. Computable trace features (word count, distinct ICD-10 codes named, and self-correction phrases) were related to reasoning ratings with Spearman correlations (exact permutation p) across the four model means and with linear mixed models (random intercept for vignette) across the 120 traces, because a feature that tracks ratings across model averages need not track individual traces. Analyses of variability across generation runs and adjudication passes were unplanned and are reported as post hoc. Analyses used R 4.5.2 (lme4, lmerTest, emmeans) and Python 3.13.1 (scipy, statsmodels).

Discordant cases

At the rating level, 18 of 486 ratings judging a model correct (3.7%) scored its reasoning 1 or below and 87 (17.9%) 2 or below; of 114 ratings judging it incorrect, 34 (29.8%) scored its reasoning 3 or above, against 399 of 486 (82.1%) of ratings judging it correct. At the trace level, averaging the five raters, no trace combined a majority-correct verdict with a mean reasoning score at or below 1.5, and 6 did so at or below 2.0. Of 23 traces the majority judged incorrect, 2 had a mean reasoning score of 3.0 or above; both concerned one vignette (169) on which raters split 2 to 3 on correctness. Ratings are clustered by vignette and rater, so these counts are reported descriptively.

**Table S5A: Pairwise comparisons of clinician-determined accuracy and clinician-rated reasoning (n = 30 vignettes)**

| Comparison | Reasoning difference (95% CI) | Holm *p* | Accuracy difference (95% CI) | Smallest equivalence margin (TOST) |
|---|---|---|---|---|
| Claude Opus 4.5 vs DeepSeek-V3.2 | +0.70 (0.50 to 0.90) | <0.001 | 0.000 (-0.071 to +0.071) | ±0.059 |
| Claude Opus 4.5 vs Gemini 3 Pro | +0.48 (0.29 to 0.67) | <0.001 | +0.007 (-0.090 to +0.104) | ±0.087 |
| Claude Opus 4.5 vs GPT-5.2 | +1.23 (1.04 to 1.43) | <0.001 | +0.060 (-0.079 to +0.199) | ±0.175 |
| DeepSeek-V3.2 vs Gemini 3 Pro | -0.22 (-0.52 to +0.08) | 0.138 | +0.007 (-0.117 to +0.130) | ±0.109 |
| DeepSeek-V3.2 vs GPT-5.2 | +0.53 (0.27 to 0.80) | <0.001 | +0.060 (-0.090 to +0.210) | ±0.184 |
| Gemini 3 Pro vs GPT-5.2 | +0.75 (0.52 to 0.99) | <0.001 | +0.053 (-0.052 to +0.159) | ±0.141 |

*Each vignette contributes one score per model, averaged over five raters; differences are paired within vignette, with 95% paired t-intervals (df = 29) and Holm-Bonferroni adjustment across the six comparisons per measure. No accuracy comparison reached Holm-adjusted $p < 0.05$. The equivalence margin is the smallest symmetric margin at which two one-sided tests pass at $\alpha = 0.05$ (the bound of the 90% interval), unadjusted; bootstrap percentile* *intervals gave the same bounds to within 0.004. After Holm adjustment, only Claude Opus 4.5 vs DeepSeek-V3.2 was equivalent at ±0.10. A crossed random-effects model (random intercepts for vignette and rater) gave identical reasoning point estimates.*

**Table S5B: Sensitivity analyses of the pairwise comparisons**

| Comparison | Unambiguous vignettes only (n = 23): reasoning | Accuracy | Excluding vignette 94 (n = 29): reasoning | Accuracy | Equivalence margin |
|---|---|---|---|---|---|
| Claude Opus 4.5 vs DeepSeek-V3.2 | +0.68 (0.45 to 0.91)* | 0.00 (−0.08 to +0.08) | +0.69 (0.49 to 0.89)* | 0.00 (−0.07 to +0.07) | ±0.061 |
| Claude Opus 4.5 vs Gemini 3 Pro | +0.56 (0.36 to 0.75)* | +0.05 (−0.05 to +0.15) | +0.48 (0.28 to 0.68)* | +0.01 (−0.09 to +0.11) | ±0.090 |
| Claude Opus 4.5 vs GPT-5.2 | +1.16 (0.97 to 1.34)* | +0.02 (−0.11 to +0.14) | +1.22 (1.02 to 1.42)* | +0.06 (−0.08 to +0.21) | ±0.182 |
| DeepSeek-V3.2 vs Gemini 3 Pro | −0.12 (−0.47 to +0.23) | +0.05 (−0.07 to +0.18) | −0.21 (−0.51 to +0.10) | +0.01 (−0.12 to +0.13) | ±0.113 |
| DeepSeek-V3.2 vs GPT-5.2 | +0.48 (0.20 to 0.76)* | +0.02 (−0.13 to +0.16) | +0.53 (0.26 to 0.81)* | +0.06 (−0.09 to +0.22) | ±0.191 |
| Gemini 3 Pro vs GPT-5.2 | +0.60 (0.36 to 0.84)* | −0.03 (−0.12 to +0.05) | +0.74 (0.50 to 0.98)* | +0.06 (−0.05 to +0.16) | ±0.146 |

** Holm-adjusted $p < 0.01$. No accuracy comparison reached Holm-adjusted $p < 0.05$ in either analysis. Unambiguous: ambiguity score 0 (Table S1C). Vignette 94 was excluded because an independent psychiatrist judged its reference diagnosis incorrect; all five raters had judged all four models correct on it. Equivalence margin: excluding vignette 94; after Holm adjustment, Claude Opus 4.5 vs DeepSeek-V3.2 remained equivalent at ±0.10 ($p = 0.028$). On the 7 ambiguous vignettes, clinician-determined accuracy was 0.67 (0.85 on unambiguous vignettes), mean reasoning 2.72 (3.08), and raters split on correctness for 8 of 28 traces (20 of 92).*

**Table S5C: Computable trace features and clinician-rated reasoning**
*A. By model (30 traces each)*

| Model | Clinician-rated reasoning | Words per trace | Distinct ICD-10 codes per trace | Traces naming no code | Self-correction phrases per trace |
|---|---|---|---|---|---|
| Claude Opus 4.5 | 3.60 | 1,088 | 2.83 | 33.3% | 0.73 |
| Gemini 3 Pro | 3.12 | 431 | 3.97 | 23.3% | 1.40 |
| DeepSeek-V3.2 | 2.90 | 620 | 3.50 | 53.3% | 3.77 |
| GPT-5.2 | 2.37 | 900 | 8.77 | 0.0% | 3.07 |

*Spearman ρ with reasoning across the four models (exact permutation p; smallest attainable two-sided $p = 0.083$): ICD-10 codes per trace −0.80 ($p = 0.33$); codes per 1,000 words −0.40 ($p = 0.75$); self-correction phrases per trace*

*−0.80 (p = 0.33); words +0.20 (p = 0.92). Of 572 code-shaped strings matched, 560 (97.9%) were chapter F codes and none were laboratory abbreviations.*

*B. Within model (120 traces; linear mixed model, reasoning ~ feature + model + (1 | vignette))*

| **Feature** | **Coefficient (95% CI)** | ***p*** | **Predicted difference across the largest between-model spread** |
|---|---|---|---|
| ICD-10 codes per trace | +0.019 (−0.007 to +0.045) | 0.145 | +0.12 |
| Self-correction phrases per trace | +0.042 (+0.009 to +0.075) | 0.014 | +0.13 |
| Codes per 1,000 words | −0.012 (−0.019 to −0.006) | <0.001 | −0.14 |
| Self-correction phrases per 1,000 words | −0.009 (−0.043 to +0.026) | 0.63 | −0.05 |
| Words, per 100 | +0.041 (+0.021 to +0.060) | <0.001 | +0.27 |

*Observed between-model reasoning differences were 0.48 to 1.23 points. Self-correction phrases were counted from a list of 14 hand-selected phrases; the raw count is length-dependent, and its density is null.*

**Section S6: Qualitative analysis**

Pipeline

We conducted a qualitative analysis of clinician commentary at multiple levels in Python (version 3.13.1). First, all comments were summarized using structured rule-based indicators reflecting the three guiding prompts (coherence, safety, flexibility) and pre-specified failure mode flags. Second, to facilitate manual thematic synthesis of free-text feedback, we analysed non-boilerplate comments containing substantive critique using semantic clustering. Specifically, comments were embedded using a sentence-level language model (“all-mpnet-base-v2”; SentenceTransformers) and grouped into a prespecified number of clusters using k-means. This clustering step was used solely as an organizational aid to surface groups of semantically similar comments and to support efficient manual review; it was not used to assign clinical meaning or determine themes. All clusters and underlying comments were manually reviewed by the study authors, who defined clinically interpretable themes and assigned each non-boilerplate comment a single primary theme based on its dominant qualitative feature. Theme prevalence was calculated separately for each model using non-boilerplate comments as the denominator.

Human judges also provided background information regarding their clinical experience, their personal and professional use of LLMs, and perspectives on LLM usage in psychiatry. Collectively, the judges had over 50 years of clinical experience, with representation across multiple subspecialties. Prior exposure to LLMs varied from weekly to daily use for professional and non-professional applications, and attitudes towards LLM use in psychiatry were generally characterized by cautious optimism regarding their potential to augment clinical practice. These background characteristics were collected to assess their potential contribution to variance in annotation behavior, rather than to support inferential comparisons between raters.

Clinician commentary flagged coherence concerns in 27.3% to 46.0% of comments per model and flexibility concerns in 1.3% to 18.0%, ordered broadly consistently with the reasoning scores (**Tables S6A** to **S6C**).

**Table S6A: Domain-level clinician concerns with model reasoning out of all comments**

| | **Coherence issues** | **Safety concerns** | **Flexibility issues** |
|---|---|---|---|
| Claude Opus 4.5 | 41 (27.3%) | 0 (0.0%) | 2 (1.3%) |
| DeepSeek-V3.2 | 55 (36.7%) | 0 (0.0%) | 8 (5.3%) |
| Gemini 3 Pro | 59 (39.3%) | 4 (2.7%) | 21 (14.0%) |
| GPT-5.2 | 69 (46.0%) | 0 (0.0%) | 27 (18.0%) |

**Table S6B: Rule-based detailed model reasoning failure modes out of all comments**

| | **Medical omission** | **No reasoning** | **Mixed coherence** | **Overflexible** | **Anchoring rigidity** |
|---|---|---|---|---|---|
| Claude Opus 4.5 | 0 (0.0%) | 0 (0.0%) | 17 (11.3%) | 0 (0.0%) | 0 (0.0%) |
| DeepSeek-V3.2 | 0 (0.0%) | 0 (0.0%) | 25 (16.7%) | 0 (0.0%) | 0 (0.0%) |
| Gemini 3 Pro | 1 (0.7%) | 3 (2.0%) | 22 (14.7%) | 1 (0.7%) | 2 (1.3%) |
| GPT-5.2 | 0 (0.0%) | 3 (2.0%) | 22 (14.7%) | 0 (0.0%) | 0 (0.0%) |

**Table S6C: Reasoning theme prevalence by model out of non-boilerplate comments**

| | Coherent but sparse reasoning | Coherent reasoning but wrong diagnosis | Post-hoc reasoning | Failure to distinguish disorder subtypes | Insufficient justification of differentials | Incoherent reasoning | Improper ranking of diagnoses |
|---|---|---|---|---|---|---|---|
| Claude Opus 4.5 | 3 (30.0%) | 1 (10.0%) | 1 (10.0%) | 2 (20.0%) | 0 (0.0%) | 1 (10.0%) | 2 (20.0%) |
| DeepSeek-V3.2 | 4 (28.6%) | 2 (14.3%) | 0 (0.0%) | 1 (7.1%) | 3 (21.4%) | 3 (21.4%) | 1 (7.1%) |
| Gemini 3 Pro | 4 (26.7%) | 4 (26.7%) | 1 (6.7%) | 2 (13.3%) | 2 (13.3%) | 1 (6.7%) | 1 (6.7%) |
| GPT-5.2 | 9 (40.9%) | 2 (9.1%) | 6 (27.3%) | 2 (9.1%) | 2 (9.1%) | 1 (4.5%) | 0 (0.0%) |

Unsafe, stigmatizing, or fabricated content

One rater flagged unsafe, stigmatizing, or fabricated content in four reasoning traces, all from Gemini 3 Pro. No other rater flagged such content for any model; one of the five raters answered only the coherence prompt in every comment. The rule-based indicator in **Table S6A** originally counted three of these four flags, because one answer (vignette 104) did not begin with "Yes"; we corrected the count by manual review. In three of the four flagged traces, all five raters judged that the model had captured the primary reference diagnosis, so these problems would not show up in accuracy scores. The comments are reproduced below verbatim from the rater's answer to the safety prompt. Reasoning scores are the mean of the five raters' diagnostic reasoning scores on the 0-4 scale.

**Vignette 94.** Reference: bipolar II disorder, current episode depressed, with high concern about suicide; unspecified anxiety disorder. Gemini 3 Pro differential: bipolar II disorder; borderline personality disorder; panic disorder; depersonalization-derealization disorder; alcohol use disorder, in early remission. Judged correct by 5 of 5 raters; mean reasoning score 3.6. Comment: "Yes, veered into the stigmatized diagnosis of borderline personality disorder on the flimsy mention of cutting and not much else."

**Vignette 104.** Reference: major depressive disorder, single episode, moderate, with psychotic features. Differential: major depressive disorder, single episode, severe with psychotic features; schizoaffective disorder, depressive type; bipolar I disorder, current episode depressed, severe with psychotic features; delusional disorder; alcohol use disorder, mild. Judged correct by 5 of 5 raters; mean reasoning score 3.2. Comment: "The jump to alcohol use disorder is incorrect and stigmatizing."

**Vignette 155.** Reference: intermittent explosive disorder. Differential: intermittent explosive disorder; narcissistic personality disorder; antisocial personality disorder; borderline personality disorder; attention-deficit/hyperactivity disorder, combined presentation. Judged correct by 5 of 5 raters; mean reasoning score 1.8. Comment: "Yes, organic causes of his symptoms (e.g. brain damage) were not explored. Also, much of the differential seems irrelevant."

**Vignette 1013.** Reference: cri-du-chat syndrome vs unspecified neurodevelopmental disorder. Differential: global developmental delay; avoidant/restrictive food intake disorder; separation anxiety disorder; unspecified neurodevelopmental disorder; unspecified mental disorder due to another medical condition. Judged correct by 0 of 5 raters; mean reasoning score 1.4. Comment: "Yes, some of the differential seemed like nonsense." In answer to the coherence prompt, the same rater wrote that the model "got to the correct diagnosis immediately, then talked itself out of it -- I think because this is a neurodevelopmental disease but the model felt pressure to find a psychiatric diagnosis."

Two of the four comments concern a stigmatizing diagnosis added to the differential on little evidence (borderline personality disorder in vignette 94, alcohol use disorder in vignette 104). The other two concern the differential as a whole: an organic cause left unexplored and irrelevant entries (vignette 155), and entries the rater judged implausible after the model set aside the correct diagnosis (vignette 1013). Because these are one rater's judgments on four traces, we report them as examples of failures that accuracy scoring misses, not as a rate.

One further comment, from a different rater on a GPT-5.2 trace (vignette 1047), noted that the schizophrenia-spectrum diagnosis in the differential "has a different treatment and can be harm[f]ul if the pt is incorrectly labelled." That rater wrote free-text comments rather than answering the three prompts, so it was not recorded as a safety flag; we list it for completeness.

**Section S7: Trainee comparison**

**Table S7: Comparison of diagnostic accuracy between Claude Opus 4.5 and two psychiatry residents on the 30 rated vignettes**

| | **Top-1 accuracy** | **Top-5 accuracy** | **Recall@5** | **Mean reciprocal rank** |
|---|---|---|---|---|
| Claude Opus 4.5 (main experiment) | 0.733 (0.556-0.858) | 0.933 (0.787-0.982) | 0.829 | 0.815 |
| Claude Opus 4.5 (this comparison only) | 0.800 (0.627-0.905) | 0.867 (0.703-0.947) | 0.776 | 0.825 |
| Resident A | 0.700 (0.521-0.833) | 0.800 (0.627-0.905) | 0.661 | 0.728 |
| Resident B | 0.500 (0.332-0.668) | 0.733 (0.556-0.858) | 0.569 | 0.599 |
| Residents pooled (60 responses) | 0.600 | 0.767 | 0.615 | 0.663 |

*Top-1 and top-5 accuracy are the majority verdict of three adjudication passes (fuzzy threshold 90) with Wilson 95% confidence intervals; recall@5 and MRR are means across passes. Reference strings that combined several diagnoses were split before scoring (**Section S4**). Residents received the vignette without the reasoning rubric supplied to models. Resident B listed fewer than five diagnoses for 11 vignettes, which limits top-5 accuracy and recall. The adjudicator declined to compare one of Resident A's first-listed diagnoses ("paraphilia", against a reference of fetishistic disorder), which was scored as incorrect; crediting it would give Resident A 0.733 and 0.833. The comparison is descriptive; no inferential claim is made.*

Reasoning was not compared between models and residents. Resident reasoning was elicited without the rubric supplied to models and is substantially sparser, and rated reasoning increases with trace length (**Table S5C**), so scoring both on the same rubric would confound reasoning quality with how it was elicited.
